# A Scoping Review of Privacy and Utility Metrics in Medical Synthetic Data

**DOI:** 10.1101/2023.11.28.23299124

**Authors:** Bayrem Kaabachi, Jérémie Despraz, Thierry Meurers, Karen Otte, Mehmed Halilovic, Bogdan Kulynych, Fabian Prasser, Jean Louis Raisaro

## Abstract

The use of synthetic data is a promising solution to facilitate the sharing and reuse of health-related data beyond its initial collection while addressing privacy concerns. However, there is still no consensus on a standardized approach for systematically evaluating the privacy and utility of synthetic data, impeding its broader adoption. In this work, we present a comprehensive review and systematization of current methods for evaluating synthetic health-related data, focusing on both privacy and utility aspects. Our findings suggest that there are a variety of methods for assessing the utility of synthetic data, but no consensus on which method is optimal in which scenario. Moreover, we found that most studies included in this review do not evaluate the privacy protection provided by synthetic data, and those that do often significantly underestimate the risks.

## Introduction

Access to high-quality data plays a crucial role in medical research and practice, particularly with the growing integration of Artificial Intelligence (AI) and Machine Learning (ML). These technologies contribute to advancements in areas like precision medicine^1^, where personalized treatments depend on comprehensive and diverse datasets. Thus, establishing safe and reliable procedures for secondary data access is important to ensure these innovations are applied ethically, securely, and effectively.

Due to privacy concerns, however, access to medical data is usually highly restricted^2^ and subject to safeguards specified in data protection laws, such as the United States *Health Insurance Portability and Accountability Act* (HIPAA)^3^ and the European Union *General Data Protection Regulation (GDPR)*^4^. A common approach used to share highly sensitive data under these regulatory frameworks is data anonymization below an acceptance re-identification risk threshold^5^. This approach employs data masking and transformation techniques to reduce privacy risks. Nonetheless, even in cases where a sufficient protection level can be achieved, anonymizing high-dimensional data often comes with a severe deterioration of the utility of the anonymized dataset,^6^ which can render it nearly unusable for research in the worst case.

A promising solution to this data-sharing problem is synthetic data, which has been described by Chen et al.^7^ as a technique that “will undoubtedly soon be used to solve pressing problems in healthcare.” The main idea behind it is to generate artificial data that mimics the statistical properties of real patient data. This data synthesis process can be achieved using multiple algorithms, including recent advancements such as *Generative Adversarial Networks*^8^ (GANs), *diffusion models*^9^, and *Large Language Models*^10^ (LLMs). These new methods generate sample that closely resemble real data, which could reduce privacy risks compared to the direct sharing of original data, and increase utility compared to anonymization.

In the medical domain in particular, several studies^11–13^ have used synthetic data to replicate case studies originally performed on real health-related data. These results highlight the potential benefits of synthetic data in the medical context and give strong arguments for the use of synthetic data as an alternative to strictly regulated personal data.

Although these results seem promising for the future of privacy-preserving data sharing in medical environments, more recent studies have pointed out risks associated with over-reliance on synthetic data as a “silver bullet” solution^14^. In particular, a malicious adversary could infer information about presence or absence of certain records in the original data, as well as infer values of sensitive attributes of known records by having access to the procedure for generating the synthetic data.^15^ This is due to the tendency of machine learning and statistical models to overfit on their training data and memorize information about individuals in the dataset^16^. Moreover, due to the black-box nature of most synthetic data generation methods such as GANs, it is difficult to predict which useful information is lost in the training-and-generation process and which sensitive information might be contained in the generated data. As a consequence, Stadler et al.^14^ argue that a cautious approach needs to be taken when generating and sharing synthetic data.

The potential risks associated with synthetic data usage highlighted in recent studies^14, 17, 18^ raise the question of whether research priorities exhibit a stronger emphasis on utility over privacy considerations. Compared to anonymized data, for which there is extensive literature^19^ describing different kinds of attacks and the corresponding privacy protection mechanisms, synthetic data has not yet been as thoroughly scrutinized. This prompted us to conduct this review in the hope of providing an informed and unbiased answer to that question.

A few surveys in the field have examined various aspects of synthetic data generation.^20, 21^ Figueira et al.^20^ provided an extensive description of multiple generation methods, and Hernandez et al.^21^ explored evaluation methods and compared them to determine the best-performing ones. In contrast to these prior studies, our approach differs in how we identify the pressing issues with synthetic data as we place a greater emphasis on the evaluation process and the privacy-utility trade-offs by having a systematic look at how synthetic data is evaluated across 73 studies. In a concurrent work, Vallevik et al.^22^ propose a taxonomy that is similar to ours in terms of fidelity, utility, and fairness. Our work, however, offers a different approach to privacy by conducting a critical analysis and comparing to the work in the Computer Science literature. We thus reach different conclusions, as we show next.

A recent series of open-source solutions such as Synthetic Data Vault,^23^ Table Evaluator,^24^ synthcity^25^ and TAPAS^15^ enable researchers to create and measure the quality of synthetic data. These platforms offer a selection of evaluation metrics and methods for assessing both utility and privacy, streamlining the evaluation process. However, these open-source tools present their own challenges as they each employ their own nomenclatures and terminologies, adding to the complexity of achieving a harmonized perspective on synthetic data within the healthcare domain. This, coupled with the presence of contradictory perspectives^14, 17, 26^ in the literature impedes the development of a unified understanding of synthetic data in healthcare.

To get a better understanding of the current landscape in healthcare-related synthetic data generation, we initiated this scoping review specifically targeting evaluation methodologies, aiming to provide a rigorous and quantitative analysis of the suitability of synthetic data evaluation methods. To do so, we have structured our analysis around answering the following two research questions:

**RQ1:** Is there consensus within the community on how to evaluate the privacy and utility of synthetic data?

**RQ2:** Is privacy and utility given the same importance when assessing synthetic data?

Synthetic medical data aims to protect patient privacy while retaining useful information. Our investigation cuts to the heart of the matter: Can practitioners trust this data to protect patient privacy and accelerate healthcare research? By investigating these two research questions, we expose the pitfalls, and provide recommendations for trustworthy synthetic data in medicine.

## Results

We reviewed articles published from 2018 to July 2024, a period that saw the rise of generative AI technologies, including the early enthusiasm in GANs and the adoption of LLMs. This growing interest is evident in our corpus as we have only two eligible publications in 2018–2019, and 21 in 2023. See Figure 1 for details.

**Figure 1.**
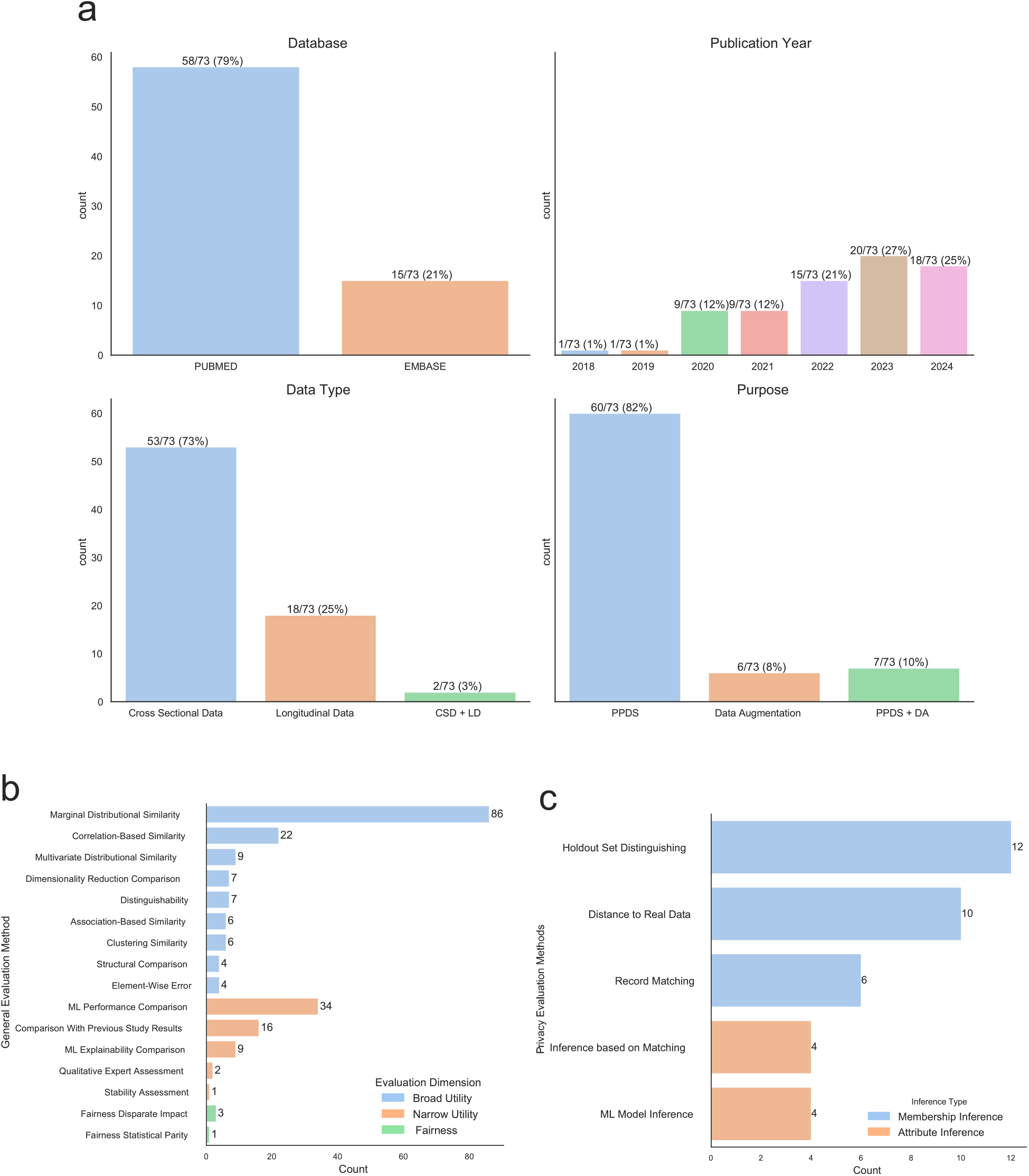
Scoping Review Results.**(a)** Visual overview of included works across various metrics. The figure depicts four dimensions: Database, Data Type, Purpose, and Publication Year. PPDS refers to Privacy Preserving Data Sharing. **(b)** Summary of the performance-related methods used in the included works. This includes a breakdown of categories such as Broad Utility, Narrow Utility and Fairness. **(c)** Summary of the performance-related methods used in the included papers. We categorized methods as Membership Inference or Attribute Inference.

After reconciling methods that were semantically the same, we found that there were 17 methods used to assess utility and 5 methods used to assess privacy. Figure 2 gives an overview of the overall landscape of utility and privacy evaluation methods used in all the publications we selected. We include the full results of the scoping review in Supplementary Data no. 1.

**Figure 2.**
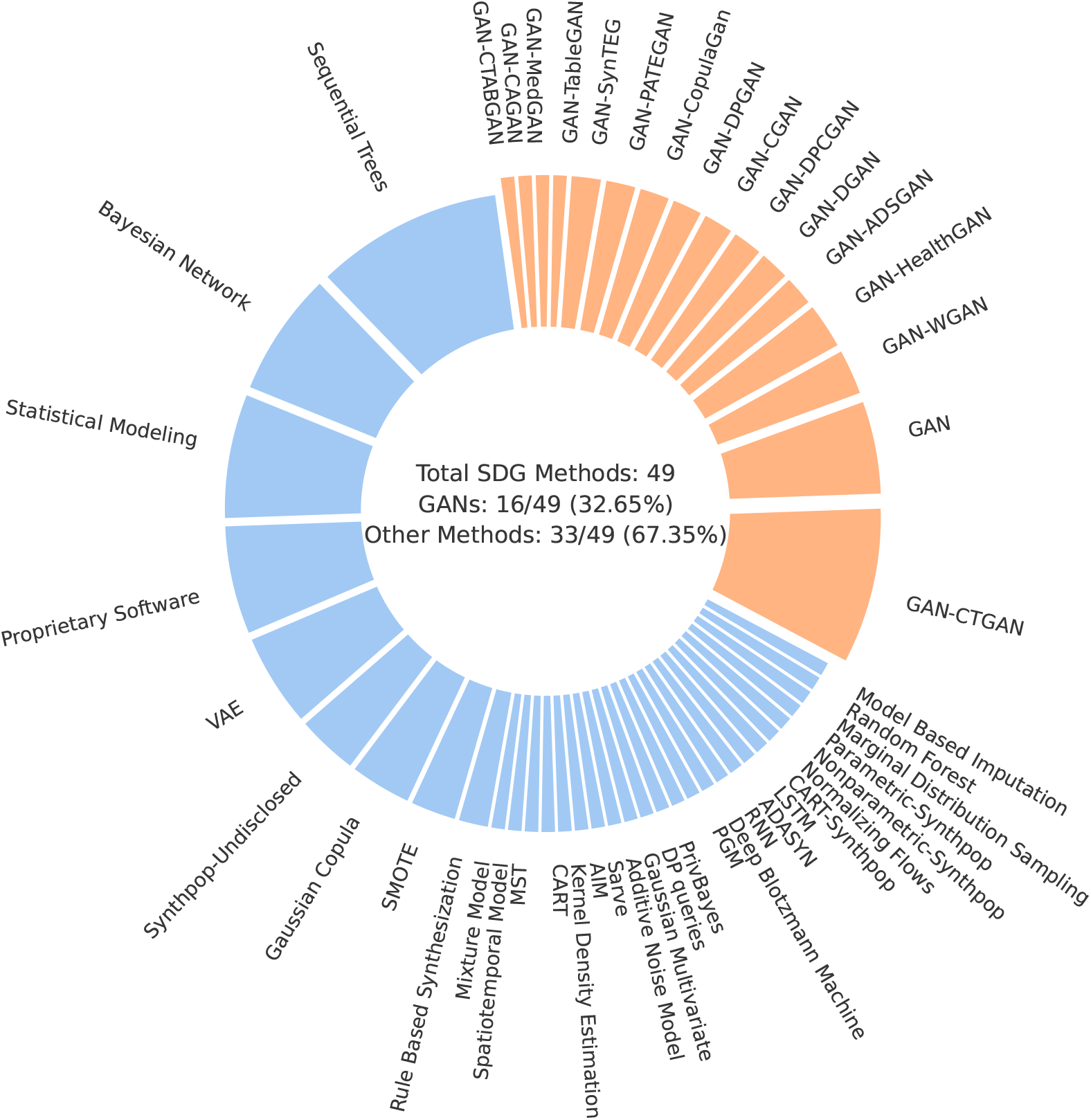
Synthetic data generation methods.We found 49 different synthetic data generation (SDG) methods, split into two main categories: GANs (Generative Adversarial Networks) and other techniques. GANs, shown in orange, make up 32.65% of the total, with 16 different methods. The remaining 67.35%, in blue, includes various approaches like Bayesian Networks, VAEs, and proprietary software.

Additionally, we found that most articles used cross-sectional data, making up 73% (53/73). Only 25% (18/73) used temporal longitudinal data, possibly as it is harder to synthesize.^27^ For this type of tabular data, the difficulty comes in maintaining relationships not just between columns which are reflected in the correlations between variables but also between rows which represent the temporal consistency of the data. As shown in Table 3, unstructured data such as images or text were not considered during this review.

**Table 1:** Key takeaways from various challenges in synthetic data generation.

| Problem | Key takeaway |
| --- | --- |
| Lack of consensus | Evaluations of synthetic data generators and synthetic data releases should cover different dimensions: broad utility/statistical fidelity, narrow utility (if synthetic data is released for a specific task), fairness, and privacy. |
| Conflicting metrics | Privacy and utility metrics that rely on similarity of synthetic records to real data, such as *nearest neighbour distance ratio*, should be used cautiously. As different studies use equivalent similarity-based metrics for measuring both utility and privacy, the usage of such metrics complicates the interpretation of the privacy-utility trade-off. |
| Principled privacy evaluation | If the purpose of synthetic data is to preserve privacy of the original data, practitioners and researchers should rigorously evaluate the associated privacy risks using modern techniques,<sup>14,15</sup> avoiding similarity-based metrics such as *distance to the closest record* or *nearest neighbour distance ratio*. |
| Ensuring provable privacy guarantees | Differential privacy (DP) is a well-established formal theory for provably guaranteeing a given level of data privacy, including for synthetic data. Although DP oftentimes can hurt utility and fairness, recent methods such as those based on $k$-way marginals<sup>36</sup> have significantly improved on its privacy-utility trade-off, making private methods a compelling candidate for synthetic data generation, especially when releasing synthetic data publicly. Even if DP is used, however, privacy risk evaluation should still be performed. |

**Table 2:** Queries by database.

| Database | Query |
| --- | --- |
| PubMed | (synthetic[Title] AND data[Title]) AND (utility[Title/Abstract] OR privacy[Title/Abstract] OR evaluation[Title/Abstract] OR metric[Title/Abstract]) |
| Embase | synthetic:ti AND data:ti AND (utility:ab OR privacy:ab OR evaluation:ab OR metric:ab) AND [2018-2024]/py |

**Table 3:** Eligibility criteria.

| Inclusion Criteria | Exclusion Criteria |
| --- | --- |
| Publications describing research that uses synthetic data generation methods and evaluates their outputs | Surveys and systematic/scoping reviews |
|  | Documents in languages other than English |
|  | Publications with no assessment of the generated output, i.e., no evaluations of the utility/privacy aspects |
|  | Publications that use unstructured data, i.e., images/text |
|  | Poster abstracts |

We found that the privacy aspect of synthetic data was the main incentive behind most selected papers with 82% (60/73) intending to use synthetic data for private data sharing scenarios. The other 8% (6/73) used it for data augmentation purposes and to answer either data scarcity or class imbalance problems. The remaining 10% (7/73) studied the potential of synthetic data in both scenarios.

Different methods were used to create synthetic data. As we show in Figure 2, out of all 49 synthetic data generation methods used in our corpus, 33% (16/49) are GANs. The rest, 67% (33/49), are a mix of other methods, including statistical modeling and methods implemented by specialized software such as Synthpop^28^ R package or the MDClone^29^ commercial platform.

Our findings indicate that the current landscape lacks a unified approach, as we identified 49 different ways to refer to utility and fairness metrics, and 22 different ways to discuss privacy which complicates the comparison and synthesis of existing evidence. We document the variability of those metrics in Supplementary Figure 1. By applying the taxonomy we proposed in Table 4 we were able to derive a trend towards *broad utility* evaluations which was noted in 153 instances (by an instance, we refer to evaluation of a specific metric, e.g., one paper can evaluate multiple metrics which all are classified as *broad utility*). *Narrow utility* is represented in 63 instances, whereas *fairness* is significantly less represented with only three instances of use. Among the works that evaluated the privacy risks of synthetic data, *membership inference* risk was the most common type, appearing in 28 instances, whereas *attribute inference* appeared in 9 instances. The specific methods used for privacy evaluation varied: 12 instances involved *holdout set distinguishing*, nine used *distance to real data*, seven employed *record matching*, five relied on *inference based on matching*, and four utilized *ML model inference*. Another notable finding is that privacy evaluations are not as often employed as utility evaluations. 95% of the studies (70/73) included utility evaluations while only 46% (31/67) of the studies claiming to employ synthetic data for preserving privacy, i.e., those that should evaluate privacy, conducted any privacy evaluation. We found that most of the studies have utilized synthetic data “as is”, assuming inherent privacy benefits without empirical verification.

**Table 4:**
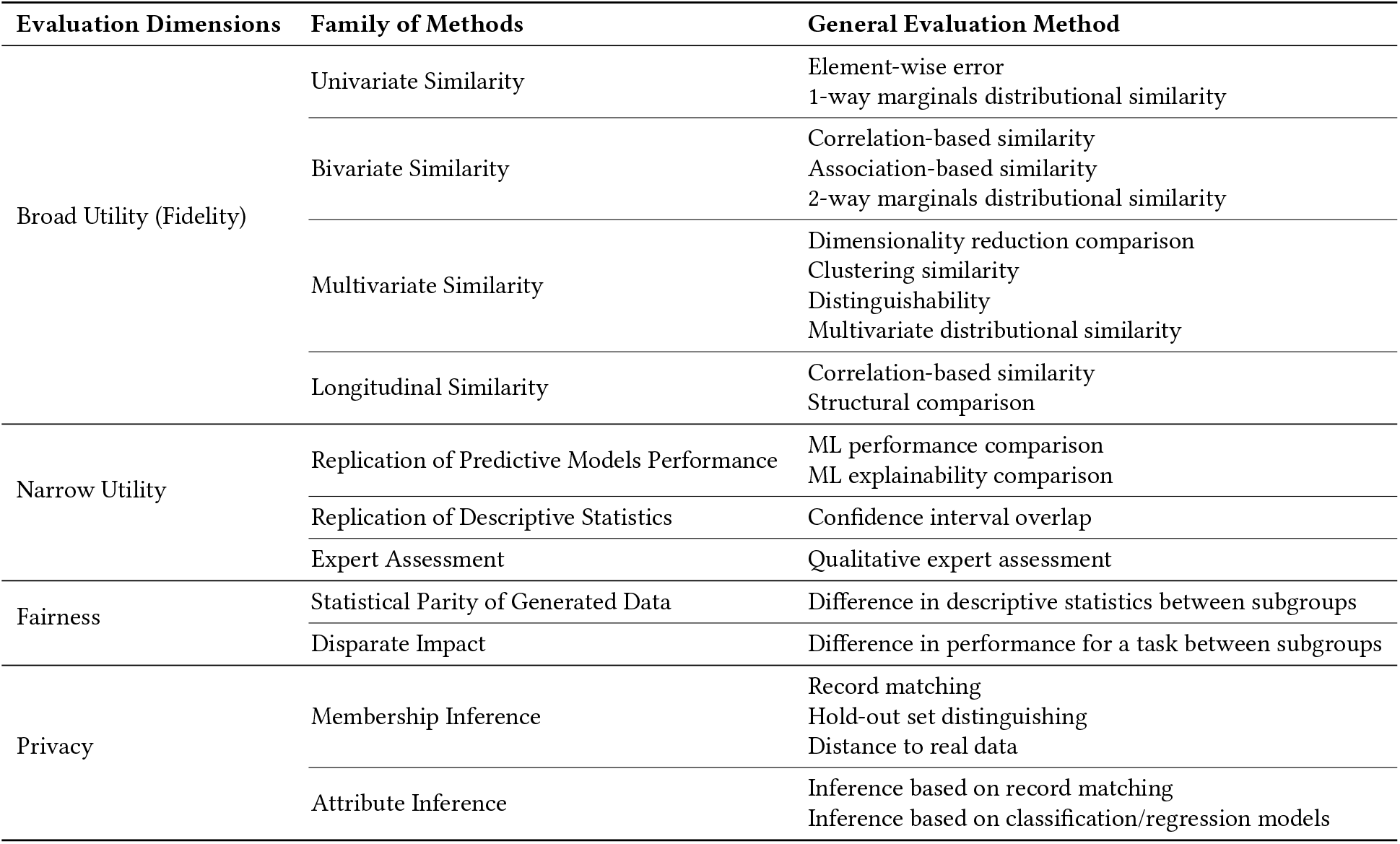
Synthetic data evaluation taxonomy.

| Evaluation Dimensions | Family of Methods | General Evaluation Method |
| --- | --- | --- |
| Broad Utility (Fidelity) | Univariate Similarity | Element-wise error<br>1-way marginals distributional similarity |
|  | Bivariate Similarity | Correlation-based similarity<br>Association-based similarity<br>2-way marginals distributional similarity |
|  | Multivariate Similarity | Dimensionality reduction comparison<br>Clustering similarity<br>Distinguishability<br>Multivariate distributional similarity |
|  | Longitudinal Similarity | Correlation-based similarity<br>Structural comparison |
| Narrow Utility | Replication of Predictive Models Performance | ML performance comparison<br>ML explainability comparison |
|  | Replication of Descriptive Statistics | Confidence interval overlap |
|  | Expert Assessment | Qualitative expert assessment |
| Fairness | Statistical Parity of Generated Data | Difference in descriptive statistics between subgroups |
|  | Disparate Impact | Difference in performance for a task between subgroups |
| Privacy | Membership Inference | Record matching<br>Hold-out set distinguishing<br>Distance to real data |
|  | Attribute Inference | Inference based on record matching<br>Inference based on classification/regression models |

In the next section, we provide a discussion of salient issues that we have identified during the analysis of these research questions, and propose concrete steps forward to rectify these issues.

## Discussion

The proposed taxonomy enables practitioners and researchers to mitigate the issue of the lack of consensus by ensuring a comprehensive evaluation within all dimensions from Table 4, covering *broad utility, narrow utility* (if synthetic data is released for a specific task), *fairness*, and *privacy*. For instance, some works in our corpus have evaluated synthetic data using multiple metrics within the same category, e.g., broad utility, yet used no metrics in other categories. Evaluating synthetic data generators or the released synthetic data across all of these dimensions provides a clearer picture of their trustworthiness.

We found that the privacy aspect of synthetic data evaluation has mostly revolved around using similarity-based metrics. It is notable that some *privacy* evaluation methods, such as *distance to real data*, can be directly at odds with equivalent metrics used for evaluating *utility*. Synthetic data is sometimes evaluated using these similarity-based metrics for both its privacy and utility even within the same study,^30^ which can lead to conflicting results and complicate the interpretation of the privacy-utility trade-off. This dichotomy highlights a challenge in harmonizing the definitions of privacy and utility in synthetic data evaluation.

The fact that most works that use synthetic data for the purpose of preserving privacy do not evaluate the residual privacy risks (see Figure 3) poses significant concern, especially with public synthetic data releases. Practitioners may inadvertently assume that synthetic data they are generating is privacy-preserving by default. This may lead to the uninformed sharing of sensitive data, potentially resulting in data breaches in addition to ethical and legal complications.

**Figure 3.**
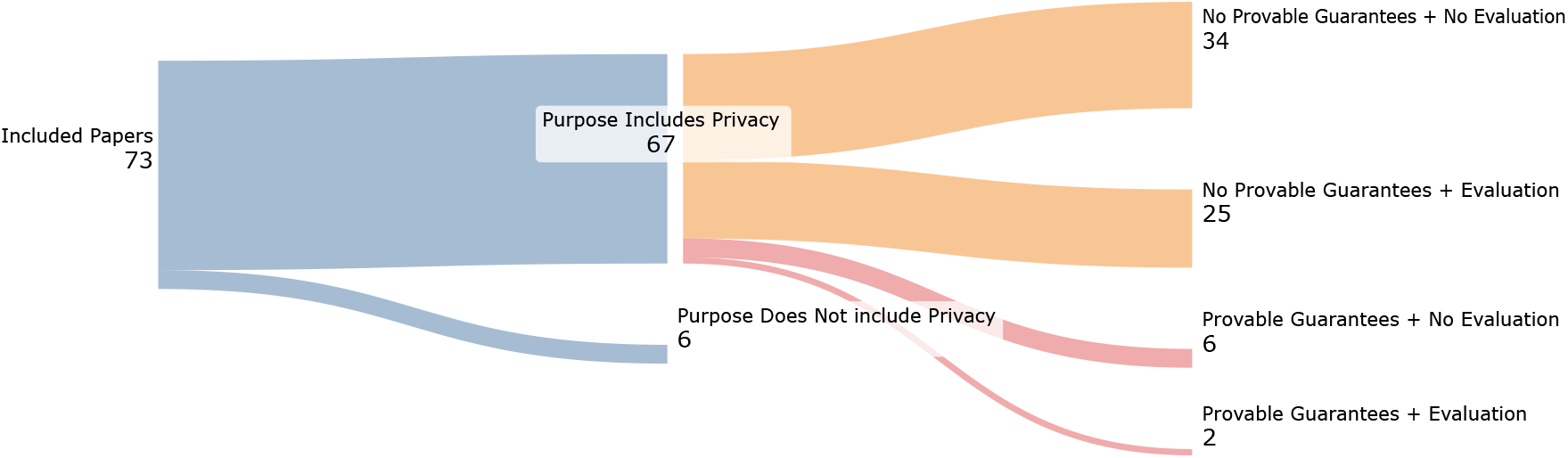
Number of works that evaluate privacy and use methods that provide privacy guarantees.Gap between the intended privacy focus of studies and the actual privacy evaluation. Only 46% (31/67) of the studies claiming to employ synthetic data for preserving privacy conducted any privacy evaluation.

Moreover, most of the studies that evaluate privacy, have employed similarity-based metrics. The prior work has recently argued and empirically demonstrated that the reliance on similarity-based metrics for privacy evaluation is inadequate for two reasons. First, such metrics do not reflect the privacy guarantees faithfully, e.g., there can exist successful inference attacks even if synthetic data is dissimilar from the original dataset.^14, 31^ Second, the publication of similarity-based metrics on its own can lead to novel privacy risks such as reconstruction attacks that leverage the reported metrics.^31^ The popularity of similarity-based metrics in our review suggests that many evaluations may offer a false sense of security regarding the privacy-preserving capabilities of synthetic data.^14^ This contrasts sharply with more sophisticated attacks discussed in the Computer Science literature, such as shadow model attacks,^14, 15^ which employ advanced techniques to assess privacy risks in a more principled way.

To ensure privacy, 11% (8/73) of the reviewed works have used differentially private^32^ synthetic data generators. Differential privacy (DP) is a well-established principled approach to ensuring provable privacy guarantees through controlled injection of random noise in the process of building the generative model. Although DP provides strong guarantees, they are significantly stronger than what is necessarily needed for practical privacy protection, which results in DP oftentimes significantly hurting utility, consistency, and fairness.^33–35^ Recent years, however, have seen significant progress in making the DP methods, including synthetic data generation, effective and feasible. In particular, there exists a family of works for generating synthetic data based on *k*-way marginals with provable guarantees both in terms of privacy and utility.^36–38^ Such methods were recently showed to be superior in terms of utility and fairness^39^ compared to other private methods based on GANs, and even, in some cases, to non-private methods.^15^ Despite this, such methods see almost no usage in the corpus we have reviewed compared to, e.g., less efficient methods based on GANs.

The level of privacy in DP is usually parameterized by the parameter *ε*, which is often criticized as non-interpretable to the practitioners.^33^ Fortunately, recent works provide operational interpretations for the level of privacy provided by DP, e.g., via success of reconstruction attacks,^40^ and argue that even if the formal privacy guarantees are weak, in practice, DP methods still provide strong resilience against practical inference attacks.^41^

Even though DP provides privacy guarantees in theory, recent studies show that practical implementations violate these guarantees due to software bugs or improper usage,^14, 42^ with a recent line of works being developed specifically to auditing the privacy guarantees afforded by DP methods.^43^ Therefore, even with theoretical guarantees, it is still important to evaluate privacy in DP synthetic data generation.

In conclusion, this review offers a detailed insight into the present research landscape of synthetic health data’s utility and privacy. The need for standardized evaluation measures stands out as a major takeaway where we believe that having uniform metrics can offer a level playing field, allowing different synthetic data generation methods to be compared in a consistent and meaningful manner. This need is increasingly apparent as international initiatives such as IEEE’s Industry Connections activity^44^ and Horizon Europe’s call for synthetic data^45^ confirm the urgency of creating clear guidelines for the development of reliable frameworks in the field. Our intention with this review is to not only shed light on these challenges, but also to inspire a collaborative effort in formulating best practices that make these techniques more accessible and understandable.

One significant concern raised throughout our work is the need for robust privacy evaluations. As the healthcare sector houses sensitive information, ensuring that synthetic data does not inadvertently lead to data leaks or result in a loss of trust is crucial. This is especially true when it comes to generative models such as GANs as their inherent complexity and lack of transparency can lead to misinformed usage where without a proper evaluation, either the privacy risks are higher than expected, or their utility is insufficient.

The integration of synthetic data in healthcare demands caution. Although it is promising, especially when using principled and provable utility and privacy-preserving methods,^36^ its potential must not be overstated. Rigorous, unbiased evaluation is crucial before implementation. Our review highlights key gaps: a lack of consensus on performance metrics, including conflicting metrics, and an absence of standardized practices for ensuring privacy guarantees. Given these shortcomings, we caution against trusting synthetic data in high-risk scenarios where false positives, missed findings, or privacy breaches could cause harm. This includes both releases for specific purposes such as medical research or decision-making, as well as public data releases. Before adopting new methods introduced in the literature or implemented in software, even those with strong guarantees, institutions should emphasize robust technical and organizational safeguards to ensure comprehensive privacy protection.

## Methods

For this scoping review, we adopted the protocol from *Preferred Reporting Items for Systematic Reviews and Meta-Analyses*^46^ (PRISMA). PRISMA stands as a recognized guideline, commonly adopted for laying out systematic reviews and meta-analyses. According to this guideline, we conduct the review by defining research questions, setting unambiguous inclusion and exclusion parameters, and detailing methods for searching, choosing, and charting data from chosen documents. We provide an overview of the procedure in Figure 4.

**Figure 4.**
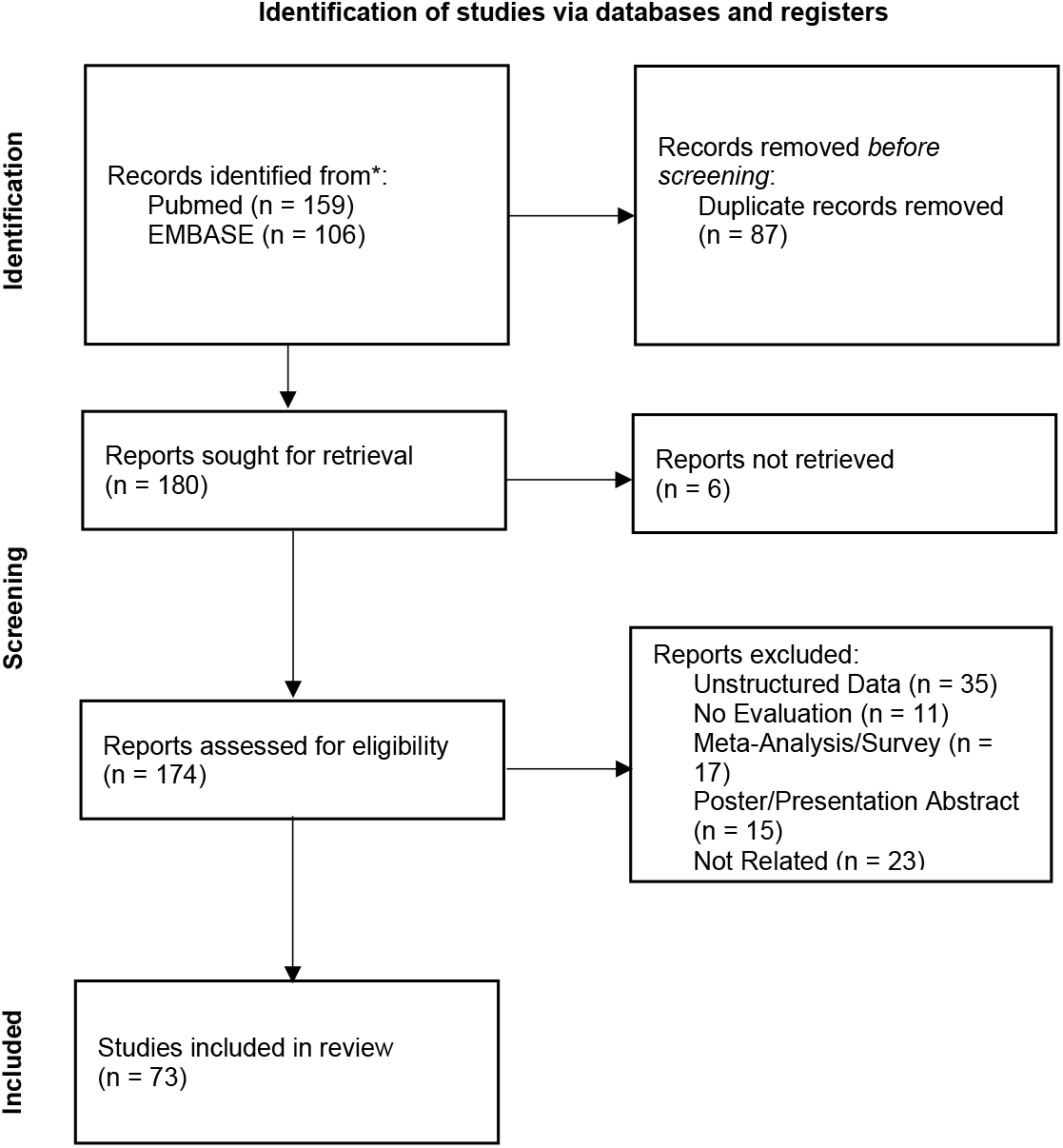
PRISMA flow diagram for the scoping review process.Identification, screening, and inclusion process of studies for the scoping review. Following the PRISMA-SCR guidelines, 174 reports were assessed for eligibility and 73 of them were included in the final review.

### Search strategy and selection criteria

To identify relevant studies, we conducted a comprehensive search across two bibliographic databases and repositories spanning the period from January 2018 to July 2024. The databases and repositories included PubMed and Embase, which are focused on healthcare and medical research. By using these biomedical databases, we could identify studies that have considered the unique constraints and requirements of healthcare settings, thus ensuring that the synthetic data methods under review would be applicable in real-world medical contexts. Full-text articles were obtained for those meeting the inclusion criteria described in Table 3. The search strategies for each database were developed at an early stage of the research and were then refined through team discussions and preliminary analysis of the results. In order to capture actionable insights on the trustworthiness of synthetic data in medicine, we designed the queries to find publications that evaluate the utility or privacy aspects of synthetic data. The queries used for each database are listed in Table 2 and were last run on July 1st, 2024.

Another consideration in query design was the avoidance of false positives, such as publications discussing synthetic compounds or materials rather than synthetic data. To this end, we included both “Title” and “Abstract” as fields for our queries, ensuring that the primary focus of the identified publications was indeed on synthetic data and its evaluation metrics for utility or privacy. We also removed such articles manually, should they have still appeared in the final selection of papers.

Any discrepancies in study selection were resolved through discussion and consensus between two of the authors. A data-charting form, illustrated in Supplementary Table 1, was collaboratively designed by the research team to delineate the specific variables to be extracted from the selected publications.

To standardize the data-charting process and ensure a unified treatment, we developed a taxonomy of evaluation methods suitable for the corpus of collected eligible publications, described next.

### Taxonomy: Performance-Related Measures

The proposed taxonomy classifies performance-related evaluation methods into three key dimensions: *broad utility, narrow utility*, and *fairness. Broad utility* (also referred to as statistical fidelity, or simply fidelity in the literature^47^) encompasses methods that we classify as *univariate similarity, bivariate similarity, multivariate similarity*, or *longitudinal similarity*. These methods are designed to capture specific aspects of data utility, ranging from straightforward one-dimensional comparisons to more complex analyses involving multiple variables and temporal patterns. This dimension is particularly valuable for making direct comparisons between different generative methods, ensuring that synthetic data can be effectively generalized across various applications and datasets. In contrast, *narrow utility* focuses on the performance of synthetic data in specific tasks or contexts. It evaluates how well the data serves particular purposes, such as improving model accuracy for a specific prediction task or supporting a specific type of statistical analysis. The *fairness* dimension examines how well synthetic data provides equitable treatment across different groups. This evaluation dimension is important as the standard measures of utility may not capture group level performance^48^ which can in turn perpetuate harmful societal biases through the use of synthetic data. We include the extended taxonomy with detailed descriptions of each family of methods and specific examples in Supplementary Table 2.

### Taxonomy: Privacy

We divide the taxonomy for privacy evaluation methods into two main categories: *membership inference* and *attribute inference*. In the *membership inference* category, we include methods which study how effectively synthetic data can prevent the identification of whether specific individuals were part of the original dataset. Based on the literature we have reviewed, this category can be subdivided into three commonly used methods: record matching, distinguishing between synthetic records and real records from a holdout set, and various techniques for computing similarity between synthetic and real data records. We classify record matching as membership inference, which is consistent with prior approaches.^49^ The *attribute inference* category addresses the risk of deducing sensitive information about individuals from synthetic data. This includes techniques like attribute inference based on record matching, which relies on conditioning on partial matches to predict specific attributes by comparing synthetic data with real data records. Another technique, *inference based on classification/regression models*, assesses how accurately private attributes can be inferred using predictive modeling approaches. As before, we provide a detailed description of the taxonomy items in Supplementary Table 2.

## Data Availability

All data produced in the present study are available upon reasonable request to the authors.

## Data Availability

The comprehensive raw dataset is included in Supplementary Data no. 1.

## Code Availability

The code utilized for data analysis is available upon request.

## Acknowledgements

Not Applicable.

## Author Contributions

Ba.K., J.D. and J.L.R. conceived the scoping review design and objectives. Ba. K. conducted database searches and screened potential articles for inclusion. J.L.R., T.M. and F.P. provided methodological guidance and critically reviewed the protocol. T.M., K.O., M.H, Bo.K. and F.P. assisted in interpreting the findings and shaping the discussion. All authors collaborated in structuring the manuscript’s narrative, Ba.K. and Bo.K. wrote the manuscript and all authors read, edited, and approved the final manuscript.

## Competing interests

The authors declare no competing interests.

## Notes

### Competing Interest Statement

The authors have declared no competing interest.

### Funding Statement

This study did not receive any funding

### Summary of Updates

Correcting Figure 3 that did not appear in the last revision. Minor typos correction.

