## Appendix for "A Scoping Review of Privacy and Utility Metrics in Medical Synthetic Data"

### Supplementary File

#### 1 Database Search Strategy

**Supplementary Table 1.** Data items used in full-text charting

| Title | Description | Possible Values |
|---|---|---|
| **DOI** | Digital Object Identifier | Free text |
| **Document Title** | Title of publication | Free text |
| **Authors** | First Author of publication | Free text |
| **Publication Year** | Year of publication | [2018..2024] |
| **Database** | Database or retrieval tool | [PubMed, Embase] |
| **General Utility Method** | General Utility Metric category | Values in Table 2. |
| **Utility Method** | Specific Utility Metric used | Values in Table 2. |
| **General Privacy Method** | General Privacy Metric category | Values in Table 2. |
| **Privacy Method** | Specific Privacy Metric used | Values in Table 2. |
| **Privacy Type** | Type of privacy involved | [Membership Inference, Attribute inference] |
| **Differential Privacy** | Use of differential privacy | [Y,N] |
| **SDG Method** | Synthetic Data Generation Method used | Free text |

#### 2 Synthetic Data Evaluation Extended Taxonomy

**Fidelity:** Refers to the accuracy with which synthetic data replicates the statistical properties and relationships of the original real data. Fidelity assessment includes various methods to determine how closely the synthetic data matches the real data across different statistical dimensions.

- **Univariate Similarity**
  - **Element-Wise Error**: Measures the difference between corresponding elements in synthetic and real datasets. Common metrics include Mean Squared Error (MSE), which calculates the average of the squared differences between corresponding data points.[1–3]
  - **Marginal Distributional Similarity**: Compares the distribution of individual variables between synthetic and real datasets to ensure that each variable's distribution in the synthetic data matches the real data. It features a multitude of different methods such as statistical tests (Mann Whitney U-test[4], T-Tests[5,6], Chi-Squared tests[5] ...),distance Between Probabilities (Wassestein Distance[7]), divergence computation (Kullback-Leibler Divergence, Hellinger Distance) and visual comparisons of marginal distributions.
- **Bivariate Similarity**
  - **Correlation-based Similarity**: Assesses the preservation of relationships between pairs of variables in synthetic data compared to real data. This method includes comparing correlation coefficients (e.g., Pearson or Spearman correlations).
  - **Association-based Similarity**: Evaluates the strength and direction of associations between variables, ensuring that the relationships in the synthetic data reflect those in the real data. This method includes Tau Statistic Comparison and Association Matrices comparison.
  - **2-Way Marginals Distributional Similarity**: Examines the joint distribution of two variables, assessing whether the synthetic data captures the correct dependencies between these variables as seen in the real data. This was usually present in the form of visual comparison of joint probabilities.
- **Multivariate Similarity**
  - **Dimensionality Reduction Comparison**: Uses techniques like Principal Component Analysis (PCA) to compare the principal components of synthetic and real data, revealing similarities or differences in underlying data structures.
  - **Clustering Similarity**: Assesses whether clusters identified in the real data are preserved in the synthetic data, using clustering algorithms to evaluate the consistency of groupings. For example, Emam et al.[7] merge synthetic data and real data, then perform a clustering algorithm. The metric is then extracted by comparing the number of synthetic samples in each cluster to the number of real samples.
  - **Distinguishability**: Measures how easily one can distinguish between synthetic and real data, often using classification tasks to determine the success rate of distinguishing the two datasets. Usually this comes in the form of ML models trained to do classification, mirroring what a Discriminator would do in a GAN architecture. Beigi et al.[8] train a classifier to distinguish between synthetic and real data then report its performance measure.
  - **Multivariate Distributional Similarity**: Examines the similarity of multivariate distributions between synthetic and real datasets, ensuring that complex interactions and dependencies among variables are accurately replicated. This is usually done with n-way marginals comparisons.
- **Longitudinal Similarity**
  - **Correlation-Based Similarity**: Evaluates the consistency of temporal correlations in longitudinal data, ensuring that trends and patterns over time are preserved in the synthetic data. This method includes comparing correlation coefficients between two time series (e.g., Pearson or Spearman correlations) or a comparison of autocorrelations.
  - **Structural Comparison**: Compares structural characteristics of data, such as trends, cycles, or other temporal features, ensuring that synthetic data accurately reflects the time-dependent structures in the real data. This method includes a comparison of directional symmetries[9] or simply visual comparisons of time series.

**Utility:** Measures the practical usefulness of synthetic data, particularly in replicating real-world scenarios and supporting decision-making processes. Utility evaluation methods ensure that synthetic data can effectively substitute real data in analytical applications.

| Evaluation Dimensions | Family of Methods | General Evaluation Method | Instances |
|---|---|---|---|
| Broad Utility (Fidelity) | Univariate Similarity | Element-wise error<br>Marginal distributional similarity | MSE<br>Marginal Distributions Visual Comparison; Kullback-Leibler Divergence; Descriptive Statistics Comparison; Wilcoxon Signed-Rank Test; T-Test; Kolmogorov-Smirnov Test; Mann Whitney U-test; Chi-Squared Test; Hellinger Distance; Wasserstein Distance; Jensen-Shannon Divergence; Distance Between Probabilities |
| | Bivariate Similarity | Correlation-based similarity<br>Association-based similarity<br>2-way marginals distributional similarity | Correlation Coefficient; Visual Comparison of Correlation Matrices; Covariance Matrix Comparison<br>Log Odds Ratio; Association Matrices Comparison; Tau Statistic |
| | Multivariate Similarity | Dimensionality reduction comparison<br>Clustering similarity<br>Distinguishability<br>Multivariate distributional similarity | PCA Visual Comparison; t-SNE Visual Comparison; FAMD Visual Comparison; Principal Components Comparison<br>Cluster Analysis<br>Distinguishability Performance<br>Maximum Mean Discrepancy; Random Projection Normality Tests; Precision; Recall; Density; Coverage; Mardia Normality Tests |
| | Longitudinal Similarity | Correlation-based similarity<br>Structural comparison | Autocorrelation Comparison<br>Visual Inspection of Distances; Directional Symmetry; Transition Matrices Comparison |
| Narrow Utility (Utility) | Replication of Predictive Models Performance | ML performance comparison<br>ML explainability comparison | ML Classification Performance; ML Regression Performance; ML RL Agent Comparison; ML Forecasting Performance Comparison<br>ML Feature Importance Comparison; ML Classification Rules Comparison; ML Feature Importance |
| | Replication of Descriptive Statistics | Confidence interval overlap | Domain Specific Metric; Replication of Studies; Comparison to Previous Study Statistics; Hazard Ratio CI Comparison; Survival Analysis Comparison |
| | Expert Assessment | Qualitative expert assessment | Domain Expert Assessment |
| Fairness | Statistical Parity of Generated Data | Difference in descriptive statistics between subgroups | Fairness Statistical Parity of Generated Data |
| | Disparate Impact | Difference in performance for a task between subgroups | Fairness Disparate Impact |
| Privacy | Membership Inference | Record matching<br>Hold-out set distinguishing<br>Distance to real data | Exact Record Match; Partial Record Match<br>Holdout Set Distance; Hypothesis Test Based on Holdout Set Distance; $\varepsilon$ -Identifiability to Holdout Set; Distance to Closest Record;<br>Nearest Neighbor Distance Ratio; Minimum Hamming Distance between Real Data and Synthetic Data; Distance to Real Data; Hamming Distance and Euclidean Distance Threshold; Cosine Distance to Real Data with Threshold; F1 Score of Model Using Distances of Synthetic Targets To Real Data; Nearest Neighbor Distance Threshold; Quantiles over Euclidean Distances to Real Data; Distance to Closest Ratio |
| | Attribute Inference | Inference based on record matching<br>Inference based on classification/regression models | Partial Matching Reconstruction; CRLProxy<br>Classification/Regression Model Attribute Prediction; Difference Between Expected and Observed Values |

**Supplementary Table 2.** Evaluation Dimensions and Corresponding Methods with Instances

- **Replication of Predictive Models Performance**
  - **ML Performance Comparison**: Assesses the performance of machine learning models trained on synthetic data compared to those trained on real data. This includes evaluating metrics like accuracy, precision, recall, and F1 score. This mostly entails classification tasks, but there have been works that compared regression performance or event reinforcement learning agents behaviour[10].
  - **ML Explainability Comparison**: Examines whether the feature importance and interpretability of models trained on synthetic data align with those trained on real data, using methods like SHAP values or feature importance scores.[11]
- **Replication of Descriptive Statistics**
  - **Comparison With Previous Study Results**: This method usually entains comparing whether the confidence intervals of key statistics in synthetic data overlap with those in real data, indicating that the synthetic data can support similar statistical analyses.[12] It also includes works that perform survival analyses on synthetic data and compare them to previously demonstrated results on real data[13].
- **Expert Assessment**
  - **Qualitative Expert Assessment**: Involves domain experts reviewing the synthetic data for its relevance, quality, and utility for specific applications, providing subjective evaluations to complement quantitative assessments.

**Fairness:** Evaluates whether synthetic data introduces or mitigates biases present in the original data. Fairness assessment is crucial to ensure equitable treatment across different demographic groups and to avoid perpetuating existing inequalities or introducing new biases.

- **Statistical Parity of Generated Data**
  - **Difference in Descriptive Statistics between Subgroups**: Assesses whether synthetic data maintains Statistical Parity of Generated Data across different demographic groups, ensuring equal representation and avoiding bias.[9]
- **Disparate Impact**
  - **Difference in Performance for a Task Between Subgroups**: Evaluates the differences in performance metrics (usually difference in True Positives) for machine learning models or other analytical tasks between different demographic groups in synthetic data, highlighting potential biases.[9]

**Privacy:** Concerns the ability of synthetic data to protect sensitive information. Privacy evaluation focuses on assessing whether synthetic data can inadvertently reveal information about individuals in the original dataset. The primary risks involve inference attacks, such as *membership inference*, where an attacker determines if an individual's data was part of the original dataset, and *attribute inference*, where sensitive attributes of individuals are deduced. These risks are particularly significant because, unlike exact data matching, synthetic data often retains statistical patterns and relationships from the original dataset, making it challenging to completely anonymize the data and prevent inference without compromising data utility. Prior works have also used adjacent terms to assess synthetic data privacy, such as *membership disclosure*. Emam et al.[14] link the two notions of inference and disclosure by describing inference as a broader type of disclosure risk.

- **Membership Inference**
  - **Record Matching**: Determines if synthetic data records can be matched to real data records, posing a risk of re-identification. This is also called Hit Rate and can take the form of a simple comparison between two records. Another way to do matching is "Partial Matching"[15], this is more relevant for cases of partial synthesis where not all attributes have been synthesized.
  - **Holdout Set Distinguishing**: Tests whether synthetic data can be distinguished from a holdout set not used in its generation, assessing overfitting and potential privacy risks.[16] Usually it has taken the form of computing a ratio such as Distance to Closest Ratio (DCR), performing hypothesis tests based on these distances, just reporting the itself. Another metric used is called CRLProxy which introduced the notion of a "trained distance" where the authors apply "a measure to calculate the distance between the representation of a known record and the representation of the synthetic records"[17].

  - **Distance to Real Data**: Uses distance metrics to measure the similarity between synthetic and real data, helping to evaluate the risk of re-identification. It has mostly taken the from of Nearest Neighbor Distance Ratio (NNDR)[18], and relied on multiple threshold based metrics such as epsilon-Identifiability to Real Data[19], threshold over quantiles, Nearest Neighbor Distance threshold, cosine distance[20], hamming distance and euclidean distance threshold[21].

- **Attribute Inference**
  - **Inference Based on Record Matching**: Assesses the risk of inferring sensitive attributes from synthetic data by matching it with real data records. Zhou et al.[22] for example perform partial matching in order to infer new information about sensitive records.
  - **Inference Based on Classification/Regression Models**: Evaluates the ability of models trained on synthetic data to predict sensitive attributes, assessing the risk of privacy breaches. Hernadez et al.[4] for example simulate attacks by providing an attacker a subset of the features. The attacker then uses ML models trained on their prior knowledge to infer the rest of the attributes.

#### 3 Extended Frequencies

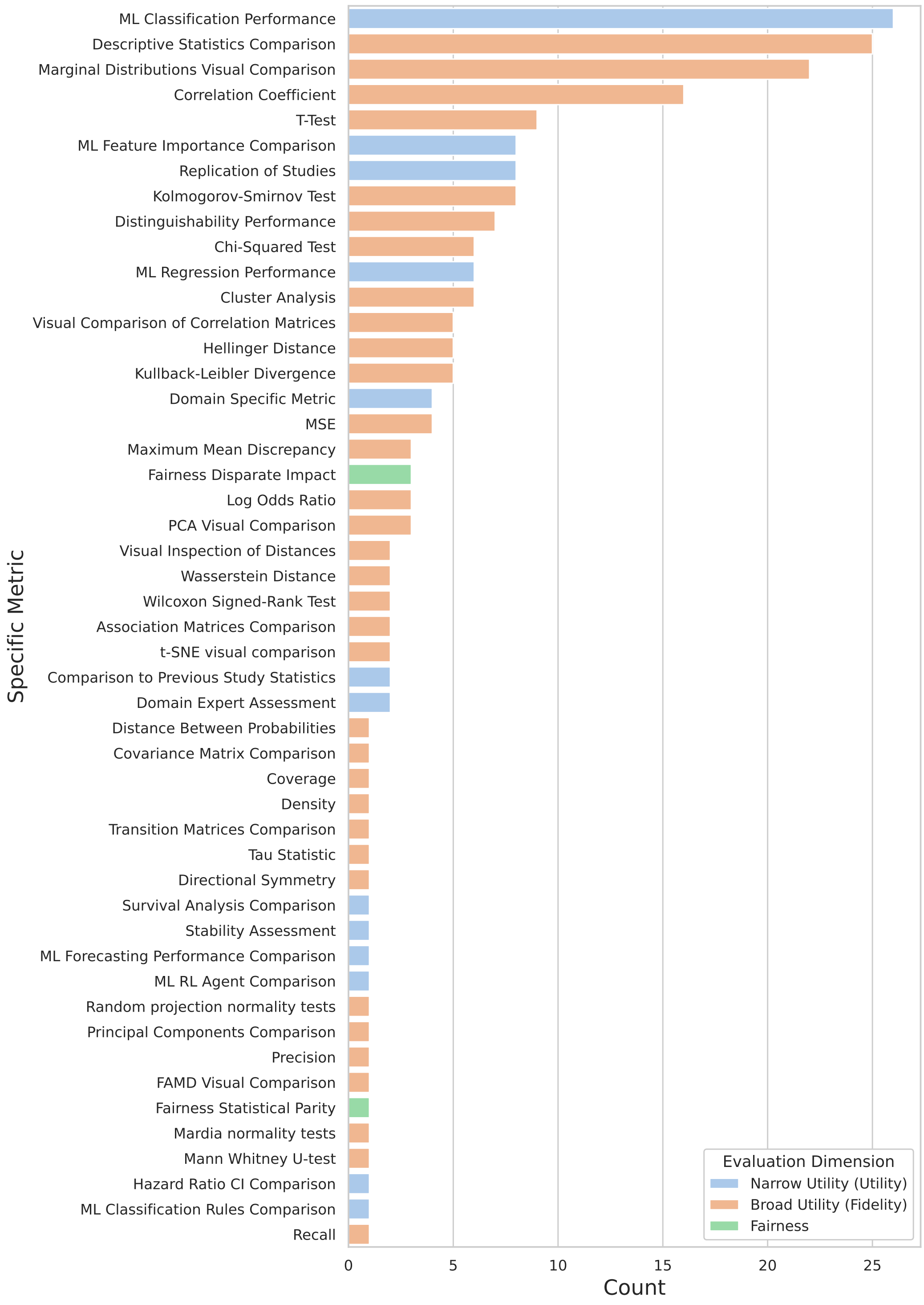


**Supplementary Figure 1.** Utility metrics frequencies. Most works evaluated synthetic data by looking at ML Classification Performance, Descriptive Statistics Comparisons and Marginal Distributions Visual Comparison.
